# Seroprevalence of anti-SARS-CoV-2 antibodies before and after implementation of anti-COVID-19 vaccination among hospital staff in Bangui, Central African Republic

**DOI:** 10.1101/2022.12.22.22283871

**Authors:** Alexandre Manirakiza, Christian Malaka, Hermione Dahlia Mossoro-Kpinde, Brice Martial Yambiyo, Christian Diamant Mossoro-Kpinde, Emmanuel Fandema, Christelle Niamathe Yakola, Rodrigue Doyama-Woza, Ida Maxime Kangale-Wando, Elliot Kosh Komba, Sandra Manuella Bénedicte Nzapali Guiagassomon, Lydie Joella-Venus de la Grace Namsenei-Dankpea, Cathy Sandra Gomelle Coti-Reckoundji, Modeste Bouhouda, Jean-Chrisostome Gody, Gérard Grésenguet, Guy Vernet, Marie-Astrid Vernet, Emmanuel Nakoune

## Abstract

**Introduction:** Healthcare workers (HCWs) are at high to very high risk for SARS-CoV-2 infection. The persistence of this pandemic worldwide has instigated the need for an investigation of the level of prevention through immunization and vaccination against SARS-CoV-2 among HCWs. The objective of our study was to evaluate any changes in anti-COVID-19 serological status before and after the vaccination campaign of health personnel in the Central African Republic.

**Method:** We carried out a repeated cross-sectional serological study on HCWs at the university hospital centers of Bangui. Blood samples were collected and tested for anti-SARS-CoV-2 IgM and IgG using the ELISA technique on blood samples.

**Results:** A total of 179 and 141 HCWs were included in the first and second surveys, respectively. Of these staff, 31.8% of HCWs were positive for anti-SARS-CoV-2 IgG in the first survey, whereas 95.7% were positive for anti-SARS-CoV-2 IgG in the second survey. However, the proportion of HCWs positive for SARS-CoV-2 IgM antibodies was low (9.7% in the first survey and 3.6% in the second survey).

**Conclusion:** These findings showed a sharp increase in seroprevalence over a one-year period. This increase is primarily due to the synergistic effect of the infection and the implementation of vaccines against COVID-19. Further studies to assess the persistence of anti-SARS-CoV-2 antibodies are needed.

## Introduction

Since the emergence of severe acute respiratory syndrome coronavirus 2 (SARS-CoV-2) in China in 2019, the world’s population has been experiencing a viral pandemic [1]. On March 11, 2020, coronavirus disease 2019 (COVID-19) was declared a pandemic disease by the World Health Organization (WHO). The WHO had reported a total of 112,456,453 confirmed COVID-19 cases and 2,497,514 deaths (7.1% case fatality rate) worldwide as of February 2021 [2].

This COVID-19 pandemic has shaken up the working world, especially in healthcare facilities. Healthcare workers (HCWs) were quickly identified as being at risk of contracting COVID-19. HCWs work around the clock and are directly involved in the diagnosis, treatment, and care of COVID-19 patients and thus are at high risk of being infected with SARS-CoV-2. In the epidemiological setting of community transmission, HCWs are also at high to very high risk for SARS-CoV-2 infection [3, 4, 5]

Although the general population needs to stay home to reduce the spread of this virus, HCWs are doing the exact opposite. In some countries, HCWs work with inadequate protection and are at constant risk of contracting COVID-19. They must be constantly monitored because, if infected, they can readily transmit the virus to their colleagues, hospitalized patients, and even family members. Increasing infection rates among HCWs could lead to the collapse of the healthcare system and a further worsening of the pandemic: if there are too few staff, the situation would be even more difficult to manage [6].

WHO has prioritized the use of vaccines and HCWs are at the highest priority level. This level of priority was motivated by the need to protect these workers to ensure the availability of critical essential services in the response to the COVID-19 pandemic and to protect them from more severe forms of COVID-19. Further, health professionals and public health authorities play a central role in discussing COVID-19 vaccination with their patients [3,7]. Epidemiological surveys can provide serological data to estimate the penetration of the virus in a given population, including the HCW population. Serological tests determine whether a person has produced antibodies in response to infection with the virus or vaccination [8].

In the Central African Republic (CAR), the Ministry of Health and Population was alerted in February 2020 and thereafter quickly implemented measures to fight the pandemic through building awareness, enhancing prevention and monitoring people traveling from high transmission areas and arriving in the CAR. On March 14, 2020, the first confirmed COVID-19-positive case was detected in the CAR. From that day until August 22, 2022, 14,803 cases have been confirmed, 14,520 patients have been cured, and 113 have died. The country has experienced four waves of this epidemic. [9, 10]. In the CAR, anti-COVID-19 vaccination has been deployed since May 20, 2021. Currently, this pandemic persists worldwide with the emergence of SARS-CoV-2 variants [11]. Accurate identification of people who have previously had COVID-19 is important in measuring disease spread and assessing the success of public health interventions [12].

Given the context, the purpose of this study was to investigate the level of prevention through immunization and vaccination against SARS-CoV-2 among HCWs in university hospitals in Bangui, the capital of the CAR. The objective of our study was to evaluate any changes in the anti-COVID-19 serological status before and after the vaccination campaign of HCWs in the CAR.

## Method

This is a repeated cross-sectional study with descriptive and analytical purposes conducted at the university hospital centers of Bangui, capital of the CAR. The first survey was conducted in April 2021 at the Pediatric University Hospital of Bangui (CHUPB), the University Hospital of the Sino-Central African Friendship (CHUASC) and the Community University Hospital (CHUC). The second survey was conducted in May 2022 at the Centre Hospitalier Universitaire Maman Elisabeth Domitien (CHUMED), the University Hospital of the Sino-Central African Friendship (CHUASC) and the Community University Hospital (CHUC) (Figure 1).

**Figure 1:**
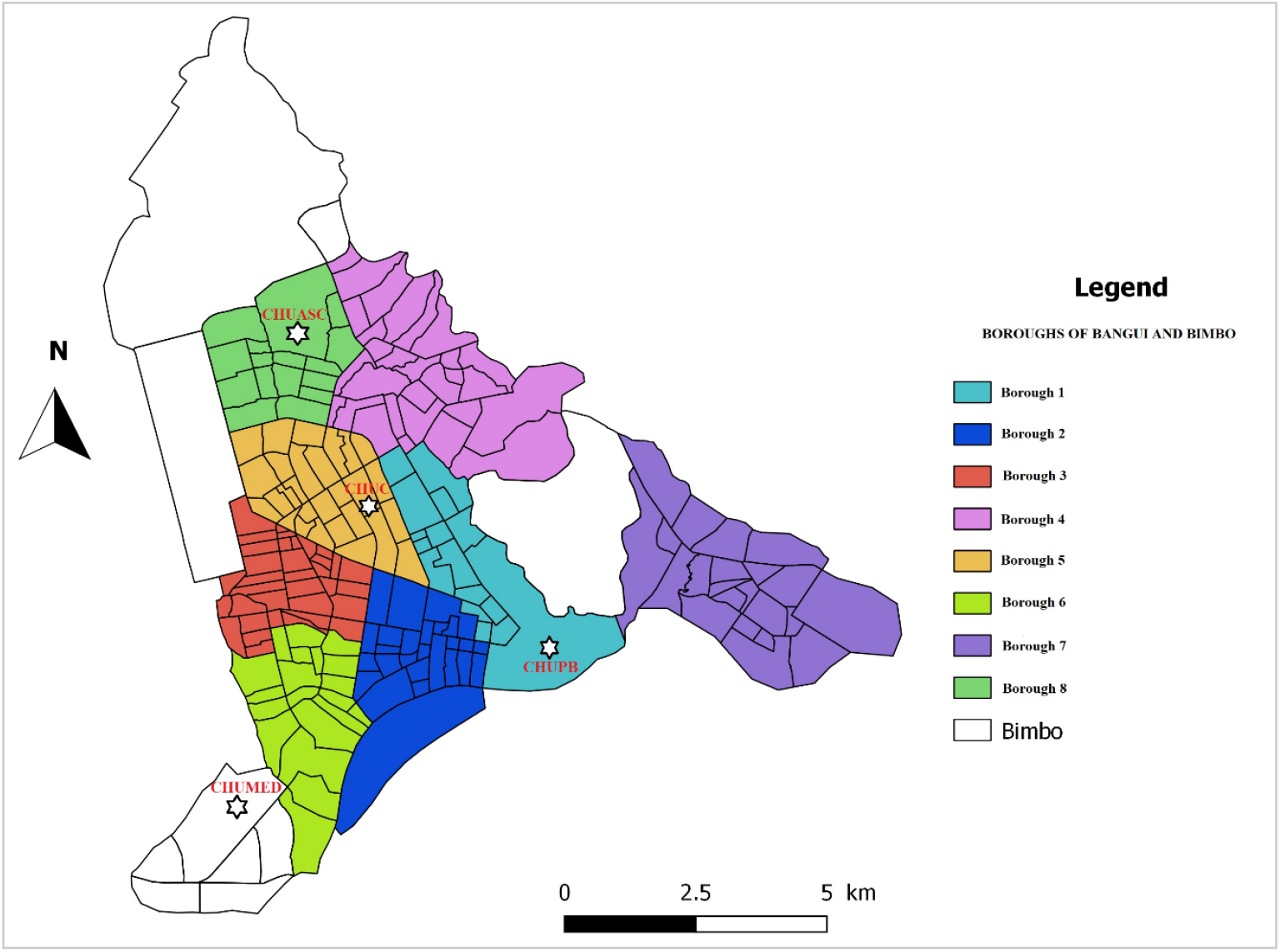
Location of hospitals included in this study (this map was created with QGIS software, https://www.qgis.org/fr/site/ from the Bangui shapefile downloaded from https://data.humdata.org/dataset/cod-ab-caf)

Our study population consisted of healthcare workers (HCWs) in hospital departments. Included were all staff administering health care to patients and support staff in the healthcare departments in the hospitals. All staff were eligible for this survey, and an informed consent form signed by each staff member was required before including them in the survey.

The protocol for this study received approval from the Ethics and Scientific Committee of the University of Bangui (N°16/UB/FACSS/CES/20) and authorization from the Ministry of Health and Population (N° 934/MSP/DIRCAB/CMPSC/20) before the survey was conducted.

The sample size for the first survey was set to 179 HCWs. The sample size was calculated using OpenEpi:

(http://www.openepi.com/SampleSize/SSPropor.htm).

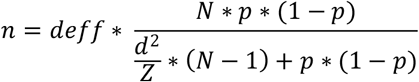

where N is the size of the population to be surveyed (for finite population correction factor) = 1000; p, the hypothesized (%) seroprevalence in the HCWs (15% +/−5 for the first survey and 15% +/−5 for the second survey; confidence limits as % of 100) (absolute +/−%) (d) = 5%; deff, a design effect (deff = 1); and Z, a constant (1.96 for a 95% confidence interval).

Based on the above parameters, the required sample size (n) was 122 participants for the second survey, for an expected seroprevalence of anti-SARS-CoV-2 antibodies equal to 90%, with a 95%confidence interval with a precision of 5%.

We collected demographic data (sex and age) and qualification of the participating HCWs.

During the first survey, nasopharyngeal swabs were collected and a blood sample of approximately 1 to 2 mL was taken in a dry tube. These samples were then sent to the Pasteur Institute of Bangui for laboratory analysis. The nasopharyngeal swabs were used for viral detection using RT-PCR (retro-transcriptase polymerase chain reaction) (polymerase chain reaction). For blood samples, serological tests were carried out to screen for anti-SARS-CoV-2 IgM and IgG using the enzyme-linked immunosorbent assay (ELISA) technique.

During the second survey, only blood samples were collected from each participant in a dry tube for serological analysis.

The data were analyzed using EpiInfo7 software. The link between the serological status (presence of IgG and IgM antibodies) of SARS-CoV-2 and the characteristics of the participating HCWs were investigated by comparing the percentages using the chi-square (χ2) test. In addition, odds ratios were calculated to assess the association between the individual characteristics according to IgM- and/or IgG-positive serological status regarding the first and second survey. Differences between groups were analyzed using a χ2 test at a P-values < 0,05.

## Results

A total of 179 and 141 HCWs were included in the first and second surveys, respectively. The proportions of socio-demographic categories such as age and gender, as well as the occupation were similar for the HCWs surveyed in both periods (P-value = 0.06, 0.24, and 0.34, respectively).

PCR analysis performed on nasopharyngeal swabs during the first survey yielded a positive result for SARS-CoV-2 in 5.9% (10/179) of the health personnel.

Given that the COVID-19 vaccine was deployed after the completion of our first survey in May 2021, during the second survey, 82.3% (116/141) of the HCWs were already vaccinated against COVID-19. Of the 116 vaccinated staff, the majority (82, or 70.8%) had received the Astra Zeneca vaccine, followed by the Johnson & Johnson vaccine (30, or 25.8%), and 4 (3.4%) had received a single dose of any of these two type of the COVID-19 vaccine.

The results of the serological analysis revealed that 51/179 (31.8%) of the HCWs were positive for anti-SARS-CoV-2 IgG in the first survey (P < 0.0001), and 95.7% of the HCWs (135/141) were positive for anti-SARS-CoV-2 IgG in the second survey. In contrast, the proportion of SARS-CoV-2 IgM-positive personnel was 9.7% in the first survey, and 3.6% in the second survey (P = 0.03). The characteristics of these two study populations are detailed in Table 1.

**Table 1:** Characteristics of the healthcare workers surveyed for seroprevalence of anti-SARS-CoV-2 antibodies in Bangui, in April 2021 (survey 1) and May 2022 (survey 2)

| Characteristics | Survey 1<br>(N=179) | Survey 2<br>(N=141) | P-value |
| --- | --- | --- | --- |
|  | n (%) | n (%) |  |
| **Gender** |  |  |  |
| Male | 49 (27.4) | 52 (36.9) | 0.06 |
| Female | 130 (72.6) | 89 (63.1) |  |
| **Mean age (years)**<br>(±SD) | 43 (± 9.7) | 43 (± 9.2) | |
| **Age category**<br>(years) |  |  |  |
| <30 | 20 (11.2) | 23 (16.3) | 0.24 |
| 31–40 | 53 (29.6) | 38 (26.9) |  |
| 41–50 | 57 (31.8) | 52 (36.9) |  |
| > 50 | 49 (27.4) | 28 (19.9) |  |
| **Hospital** |  |  |  |
| CHUPB | 40 (22.4) | - | NA |
| CHUMED | 57 (31.8) | 61 (43.3) |  |
| CHUC | 82 (45.8) | 55 (24.8) |  |
| CHUASC | - | 45 (31.9) |  |
| **Occupation** |  |  |  |
| Doctors | 36 (20.1) | 31 (22.0) | 0.34 |
| Nurses and<br>midwives | 52 (29.1) | 49 (34.7) |  |
| Other | 93 (50.8) | 61 (43.3) |  |
| **Anti-COVID-19**<br>**vaccination** |  |  |  |
| Yes | * | 116 (82.3) | NA |
| No | * | 25 (17.7) |  |
| **PCR** |  |  |  |
| Positive | 10 (5.6) | * | NA |
| Negative | 169 (94.4) | * |  |
| **Anti-SARS-CoV-2**<br>**IgG** |  |  |  |
| Positive | 51 (31.8) | 135 (95.7) | <0.0001 |
| Negative | 122 (68.2) | 6 (4.3) |  |
| **Anti-SARS-CoV-2**<br>**IgM** |  |  |  |
| Positive | 17 (9.7) | 5 (3.6) | 0.03 |
| Negative | 162 (90.5) | 136 (96.4) |  |

CHUPB, Pediatric University Hospital of Bangui; CHUMED, Centre Hospitalier
Universitaire Maman Elisabeth Domitien; CHUC, Community University Hospital;
CHUASC, the University Hospital of the Sino-Central African Friendship

The analysis of the relationship between the characteristics of the participating HCWs, neither gender (male or female), nor age categories had any effect on the positive serological status (IgM and/or IgG) in the two surveys. In the hospitals where we conducted the two surveys successively (CHUC and CHUASC), the positive serological status increased from 58.5% during the first survey to 100% during the second survey in the CHUC, whereas it increased from 77.2% to 96.7% in the CHUASC (OR, 1.83; 95% confidence interval, [1.02-3.3]; P = 0.041). Regarding the Occupation of the HCWs surveyed, personnel not directly involved in health care were less likely to have a positive serological status (IgM and/or IgG) against SARS-CoV-2 compared with medical doctors ([0.26-0.90]; P = 0.02), but this risk was similar between doctors and nurses and midwives (Table 2).

**Table 2:** Relationship between healthcare workers and COVID-19 serological status (IgM and/or IgG) in Bangui, April 2021 (survey 1) and May 2022 (survey 2)

| Characteristics |  | Survey 1 (N=179) |  | Survey 2: (N=141) |  | OR* | [95%IC] | P-value |
| --- | --- | --- | --- | --- | --- | --- | --- | --- |
|  |  | Positive IgM and/or IgG: n (%) | Negative IgM and/or IgG: n (%) | Positive IgM and/or IgG: n (%) | Negative IgM and/or IgG: n (%) |  |  |  |
| **Gender** |  |  |  |  |  |  |  |  |
|  | Male | 20 (40.8) | 29 (59.2) | 50 (96.2) |  | Ref. |  |  |
|  | Female | 45 (34.6) | 85 (65.4) | 85 (95.5) |  | 0.75 | [0.40-1.42] | 0.38 |
| **Age category (years)** |  |  |  |  |  |  |  |  |
|  | ≤30 | 14 (70.0) | 6 (30.0) | 21 (91.1) | 2 (8.7) | Ref. |  |  |
|  | 31 à 40 | 31 (58,5) | 22 (41,5) | 36 (94,7) | 2 (5,3) | 0,77 | [0.34-1.77] | 0,54 |
|  | 41 à 50 | 38 (66,7) | 19 (33,3) | 50 (96,2) | 2 (3,8) | 1,07 | [0.48-2.42] | 0,86 |
|  | > 50 | 31 (63,3) | 18 (45,0) | 28 (100,0) | 0 (0,0) | 0,60 | [0.26-1.40] | 0,24 |
| **Hospital** |  |  |  |  |  |  |  |  |
|  | CHUPB | 22 (55.0) | 18 (45.0) | - | - | NA |  |  |
|  | CHUMED | - | - | 41 (91.1) | 4 (8.9) | NA |  |  |
|  | CHUC | 49 (58.5) | 34 (41.5) | 35 (100.0) | 0 (0.0) | Ref. |  |  |
|  | CHUASC | 44 (77.2) | 13 (22.8) | 59 (96.7) | 2 (3.3) | **1.83** | [1.02-3.3] | 0.041 |
| **Occupation** |  |  |  |  |  |  |  |  |
|  | Doctors | 26 (72.2) | 10 (27.8) | 28 (90.3) | 3 (9.7) | Ref. |  |  |
|  | Nurses and midwives | 36 (69.2) | 16 (30.8) | 48 (98.0) | 1 (2.0) | 1.24 | [0.63-2.46] | 0.54 |
|  | Other | 114 (63.7) | 65 (36.3) | 59 (96.7) | 2 (3.3) | **0.45** | [0.26-0.90] | 0.02 |

\*OR, odds ratio calculated by comparing IgM and/or IgG positive serological status between the first and second survey. Significant values are
shown in bold. CHUPB, Pediatric University Hospital of Bangui; CHUMED, Centre Hospitalier Universitaire Maman Elisabeth Domitien;
CHUC, Community University Hospital; CHUASC, the University Hospital of the Sino-Central African Friendship

## Discussion

The results of this study on the seroprevalence of anti-SARS-CoV-2 antibodies are the first data on the serological status of COVID-19 among HCWs in the CAR. Here, we assessed this indicator over two different periods, revealing a significant increase in the proportion of HCWs with anti-COVID-19 antibodies after one year. This is due to the continuous circulation of the virus in the general population, given that these HCWs can not only become infected in the community where they live, but they are also relatively more exposed to infection in their workplaces. Our study is in line with other studies monitoring the seroprevalence of anti-SARS-CoV-2 antibodies and the acquisition of anti-SARS-CoV-2 immunity in HCWs. Similar studies on this topic have been carried out in several countries since the emergence of the COVID-19 pandemic with varied results [13, 14, 15, 16]. Many national and regional studies have estimated the prevalence of SARS-CoV-2 IgG antibodies in the population, and the seroprevalence in Africa (65.1% in Q3 2021) is among the highest in the world [17, 18]. A recent sero-epidemiological survey conducted in Bangui showed that there is a high cumulative level of immunity in general population, thus indicating a significant degree of spread of SARS-CoV-2 in the population [13].

The synergistic effect of the COVID-19 infection and the vaccine that the staff received explains this high level of seroprevalence. Although the seroprevalence found during the first survey of this study was driven only by SARS-CoV-2 infection, the sharp increase in seroprevalence observed with the second survey indicates a complementary increase in immune status through vaccination. This sharp increase in seroprevalence may be due also to the infections caused by more transmissible variants [19].

The IgG level in the vaccinated population can remain elevated for 25 weeks, indicating that IgG may exist for an even longer period with a positive effect against SARS-CoV-2 [17]. Our IgM and IgG antibody seropositivity rates corroborate those from other countries. For example, in a study of IgG and IgM response to SARS-CoV-2 vaccine in HCWs in China in July 2021, IgM and IgG antibody seropositivity rates were 3.1% and 74.2%, respectively [18].

## Conclusion

Our findings showed us that almost all HCWs in Bangui had developed IgG antibodies to SARS-CoV-2 by May 2021. This result confirms the effect of vaccination as well as infection with SARS-CoV-2, because this virus has been circulating intensively in the general population. However, further studies are now needed on the persistence of neutralizing antibodies, the enhancement of immunogenicity against SARS-CoV-2 variants and the impact of vaccination based on timely seroprevalence data.

## Data Availability

Data underlying our findings are made fully available without restriction

## Author contributions

Conceptualization and methodology: AM, CM, GV, GG, EN and MAV; field investigation: HDMP, CDMP, EKK; RHD, CNY, and EF; supervision: AM, IMK, SMBNG, CSGR, JCG and MAV; laboratory analysis: MAV, CM and MB; data verification and analysis: MAV, CM, BY, LJN and MA; draft preparation and writing: all authors

## Acknowledgments

We thank the healthcare workers who participated in this study. We express our gratitude to the hospital administration authorities for their help and advice in collecting the data. Many thanks also to Vincent Richard of the International Affairs Department at the Pasteur Institute of Paris for providing support for this study.

## Declaration of Competing Interest

All authors declare no competing interests.

## Funder

This study was financially supported by the European Union via MEDILABSECURE (https://www.medilabsecure.com/) and the Institut Pasteur Network association. The funder of the study had no role in study design, data collection, data analysis, data interpretation, or writing of the report.

## Data Availability

All supporting data are available from the corresponding author on reasonable request

## Notes

### Competing Interest Statement

The authors have declared no competing interest.

